# The Risk factors for fear of falling in chronic stroke patients: a systematic review and meta-analysis

**DOI:** 10.1101/2023.11.01.23297920

**Authors:** Yun Kong, Kelong Zhong, Xuemei An

## Abstract

**Background:** The incidence rate of fear of falling in chronic stroke is high, which seriously affects the quality of life and rehabilitation effect of patients. Early identification of its risk factors will help clinical screening of high-risk patients and prevent their further development. There is currently no systematic evaluation of risk factors for fear of falling falls in chronic stroke.

**Objective:** We systematically reviewed the literature on risk factors for fear of fall in chronic patients with stroke.

**Method:** We systematically searched PubMed, Embase,cochrane,Web of Science and the China National Knowledge Infrastructure(CNKI),the china biomedical literature database(CBM),the China Science and Technology Periodicals Database(VIP),Wangfang data for relevant literature until May 2023.Review Manager V.5.3 merged the OR value and 95% CI of the potential risk factors.A random/fixed-effect meta-analysis was used to pool risk factors from individual studies.Cochran’s Q and the I^2^ tests were used to assess heterogeneity between the studies.

**Results:** A total of 6 studies were included for the final analysis, with 965 chronic stroke patients. The risk factors for fear of falling in chronic patients with stroke were impaired balance ability (OR=3.05; 95% CI 1.60 to 5.80), history of falls (OR=2.12; 95% CI 1.40 to 3.20) and anxiety (OR*=*2.29; 95% CI 1.43 to 3.67), depression (OR=1.80; 95% CI 1.22 to 2.67), poor lower limb motor function (*OR*=1.14; 95% CI 1.00 to 1.29),physically inactiveness (OR=2.04; 95% CI 1.01 to 4.12). Married (OR=0.61; 95% CI: 0.435-0.875) is a protective factor.

**Conclusion:** Our study shows that impaired balance ability, history of falls might be a greater risk for fear of falling. Future studies are recommended to determine other risk factors specific to patients with chronic stroke.

---

Stroke remains the second largest cause of death globally and the main cause of disability in adults(1). In China, stroke is the main cause of death, and As our population grows and ages, the number of deaths will steadily increase(2).There are many complications after brain stroke, and falls are a common complication(3).Some scholars have found that people with a history of stroke have an increased risk of falls (4),which may be related to hemiplegia(5), balance loss(6), sensory disturbances(7), fatigue(8), depression and anxiety(9), and cognitive dysfunction(10) caused by stroke.Fear of falling (FOF) refers to the decrease in self-efficacy or confidence that occurs to avoid falling while performing an activity(11).Studies have shown that patients with stroke have a higher risk of developing the fear of falling(12), which may be related to an increased risk of falling after stroke(13).The incidence of fear of falling after stroke has been reported to be approximately 32 to 66 percent(14–16). Fear of falling is associated with adverse consequences such as limited activity, increased incidence of falls, decreased quality of life,and increased hospitalization and case fatality rates(17, 18).Moreover, FOF was also significantly associated with anxiety and depression(16).Therefore, it is very important to screen stroke people with high risk of fear of falling as early as possible and provide targeted intervention measures, so it is necessary to explore the risk factors of FOF in stroke.However, the fear of falling is a dynamically changing psychological problem, and risk factors vary from stage to stage.Previous studies have always considered acute stroke and chronic stroke as a whole, so this study only conducts a meta-analysis of risk factors for fear of falling in patients with chronic stroke, in order to provide a reference for clinical staff to identify people at high risk of FoF as early as possible and prevent their further development.

## 1. Materials and methods

### 1.1 Search Policy

Using subject words combined with free words, we searched the CNKI, Wanfang, VIP, CBM, PubMed, EMbase, The Cochrane Library, and web of science databases. The search period is until May 30, 2023.search terms include: “stroke”, “strokes”, “cerebrovascular accident *”, “CVA”, “cerebrovascular apoplexy”, “brain vascular accident *”, “cerebrovascular stroke *,” apoplexy “,” cerebral stroke, “acute stroke,” acute cerebrovascular accident “,” cerebral infarction “,” cerebral hemorrhage “,” brain ischemia, “Accidental Falls”, “Falls”, “Falling”, “Accidental Fall*”, “Slip and Fall”, “Fall and Slip”, “Accident Prevention “,” Home Accidents “,” Fear “,” fear* “,” Panic “,” worry “,” afraid”. Taking the PubMed database as an example, the retrieval formula is as follows:

(((stroke[MeSH Terms]) OR (“stroke”[Title/Abstract] OR “strokes”[Title/Abstract] OR “cerebrovascular accident*”[Title/Abstract] OR “CVA”[Title/Abstract] OR “cerebrovascular apoplexy”[Title/Abstract] OR “brain vascular accident*”[Title/Abstract] OR “cerebrovascular stroke*”[Title/Abstract] OR “apoplexy”[Title/Abstract] OR “cerebral stroke*”[Title/Abstract] OR “acute stroke*”[Title/Abstract] OR “acute cerebrovascular accident*”[Title/Abstract] OR “cerebral infarction”[Title/Abstract] OR “cerebral hemorrhage”[Title/Abstract] OR “brain ischemia”[Title/Abstract])) AND ((Accidental Falls[MeSH Terms]) OR (“Falls”OR“Falling”OR “Accidental Fall*”OR “Slip and Fall”OR “Fall and Slip”OR “Accident Prevention”OR “Home Accidents”[Title/Abstract]))) AND ((Fear[MeSH Terms]) OR (“fear*”[Title/Abstract] OR “Panic”OR “worry”OR“afraid”[Title/Abstract])).

### 1.2 Inclusion and exclusion criteria

Inclusion criteria (1) Study subjects: stroke patients who meet the diagnostic criteria, patients with disease time ≥3 months or convalescent period; (2) exposure factors: risk factors for fear of falling in patients with chronic stroke were reported, and the OR or RR value was extracted directly from the included studies; (3) fear of falling: use of validated screening tools; (4) Case-control studies, cohort studies, and cross-sectional studies.

Exclusion criteria: (1) the data in the literature cannot be converted and applied; (2) repeated publications; (3) Conference papers, case reports; (4) low-quality literature.

### 1.3 Literature screening and data extraction

Endnote X9 software was used to remove duplicates.Browsing the title and abstract, preliminary screening of literature according to inclusion and exclusion criteria, further review of literature that may meet the inclusion criteria, discussion or consultation with a third person in case of disagreement.Two review authors conducted literature extraction independently based on searches, reviewed and selected according to predefined criteria.The extracted data includes:first author, year of publication,Country, type of study, sample size, course of illness, screening tools for fear of falling, influencing factors.

### 1.4 Quality assessment

Two researchers independently evaluated the literature, and if there was a disagreement, a third investigator was invited to participate.The methodologic quality assessment of case–control studies and cohort studies was assessed by the Newcastle Ottawa Scale (NOS) for the study population,comparability and outcome evaluation. The scale’s total score was kept as 9 points, where 0 to 3 were divided into low-quality research, 4 to 6 were divided into medium-quality research and 7–9 were divided into high-quality research.In addition, the risk of bias in a cross-sectional study was assessed using the instrument Agency for Healthcare Research and Quality (AHRQ).The total score is 11 points, 0∼3 is low quality, 4∼7 is medium quality, and 8∼11 is high quality.

### 1.5 Statistical analysis

we conducted a meta-analysis by the RevMan V.5.3 software to pool the *OR/RR* value with 95% CI.Statistical heterogeneity between studies was quantified by the I^2^ statistics and formally tested by Cochran’s Q statistic. If P≥0.1, I^2^≤50%, indicating homogeneity between studies, a fixed-effect model was used; Conversely, there was heterogeneity, and random-effects models were used or results pooled were abandoned.Sensitivity analysis: different effect models were used to pool effect sizes separately to see if there was a difference in the results.

## 2 Results

### 2.1 Literature selection

Initially,1297 records were searched from the eight databases and other resources (figure 1). After the exclusion of duplicates, the remaining 669 records were screened. After analysing the title and abstract, ultimately, 43 publications were selected for the full-text assessment. Finally, six full-text studies were included in this meta-analysis.

**Fig 1.**
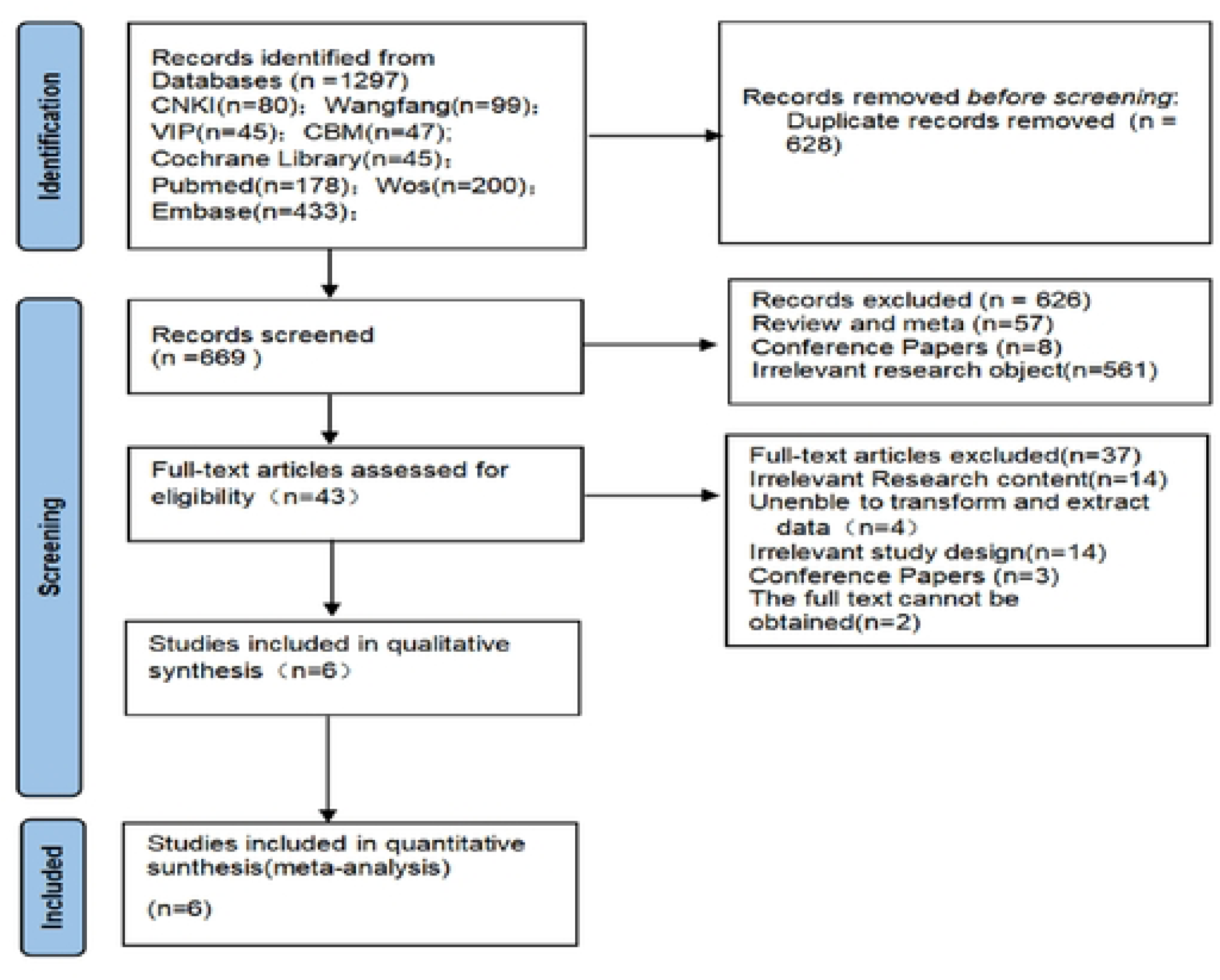
Flow chart of literature screening.

### 2.2 Basic characteristics and quality evaluation of the included literature

Six studies were included in the meta-analysis.They are from four countries: China (n=2), Sweden (n=2), Canada (n=1), India (n=1);Among these studies, two were cross-sectional, three were case–control and one were prospective cohort studies. the included literature scored as 5-8; 4 high-quality studies and 2 of moderate quality. A summary of literature characteristics used in the analysis is shown in table 1.

**Table 1.**
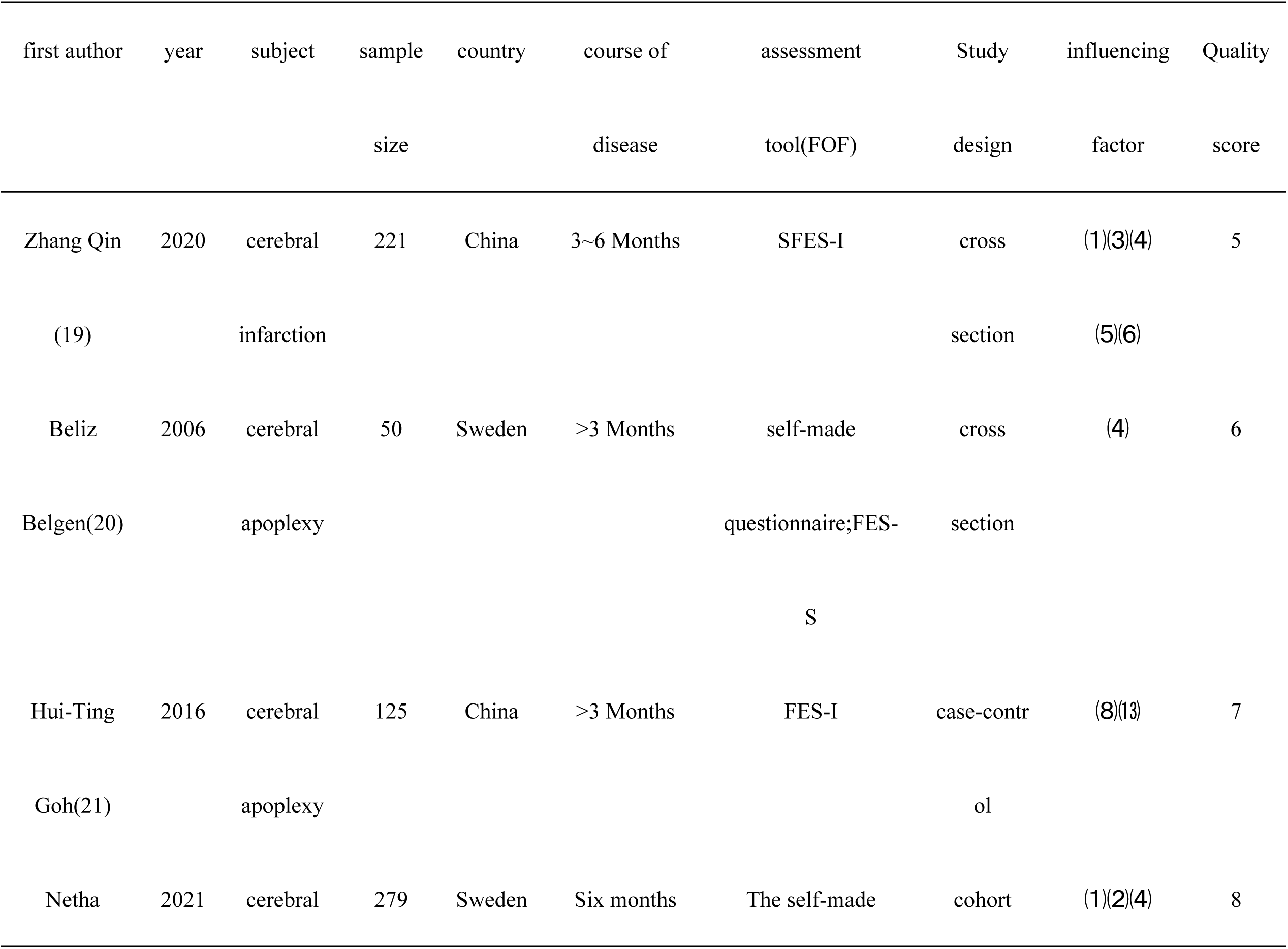
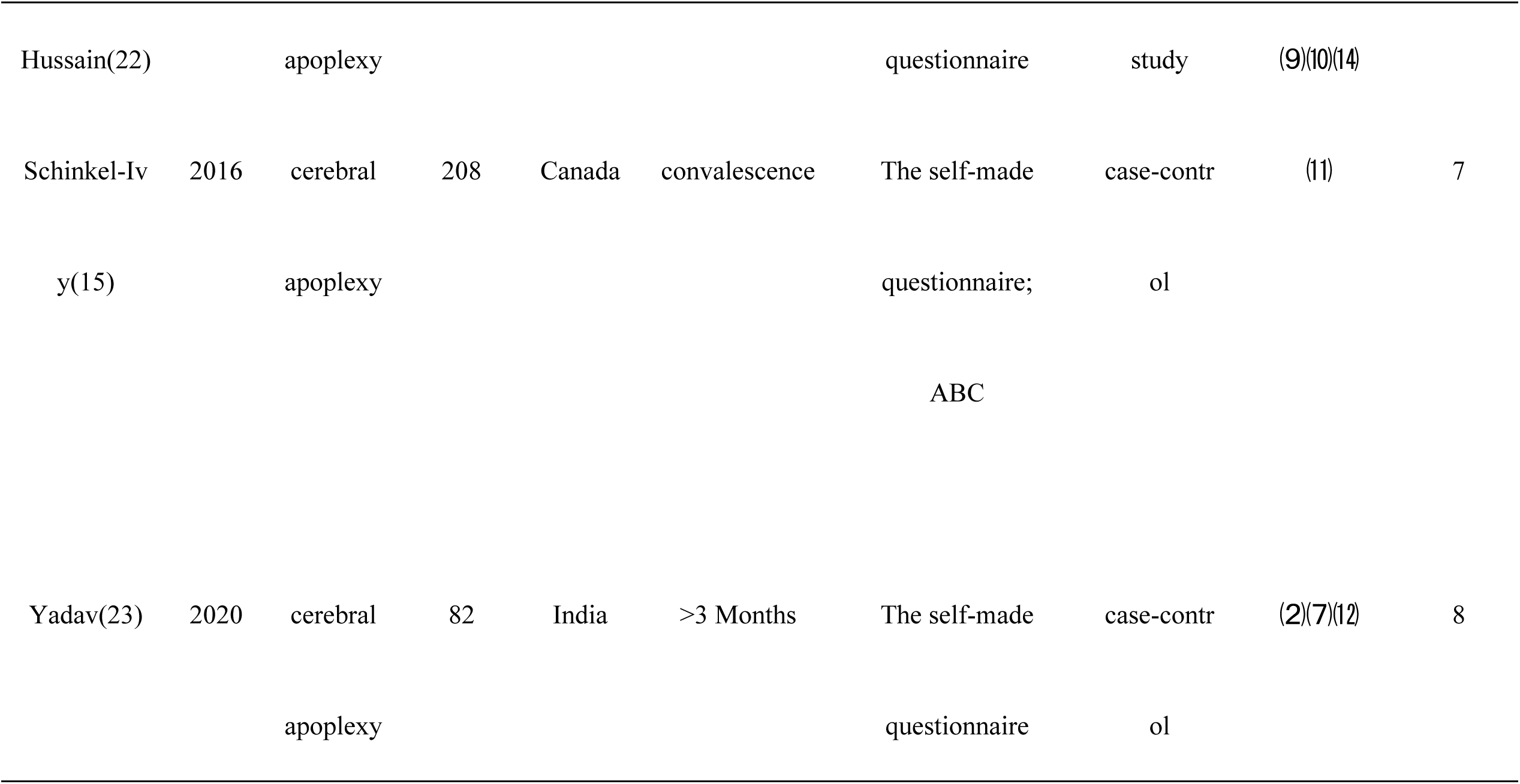
has included the basic characteristics of the literature.

### 2.3 Results of the meta-analysis

#### 2.3.1 Age

Two studies reported the relationship between age and fear of falling, with heterogeneity testing (I2=82%, p=0.02), with great heterogeneity among the studies, so a random effects model was used for the analysis. The results showed no statistical significance:OR=1.13;95% CI:0.85-1.52(Figure 2).

**Fig 2.**
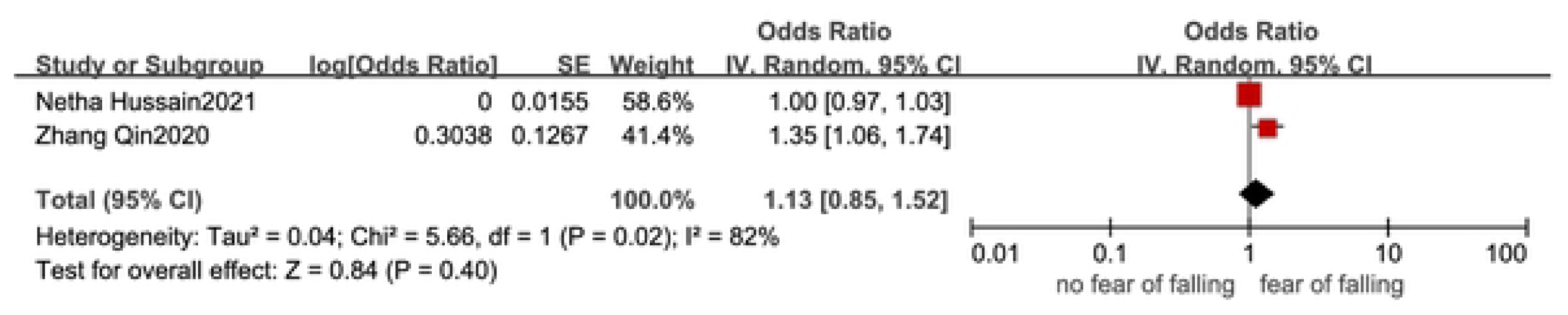
Forest plot of the relationship between age and fear of falling.

#### 2.3.2 History of falls

Three studies reported the relationship between history of falls and fear of falling, with heterogeneity testing (I2=0%, p=0.93), and there was no heterogeneity among the studies, so a fixed-effects model was used for the analysis. The results showed that history of falls (OR=2.12; 95% CI:1.40-3.20) was a risk factor for fear of falls in chronic stroke(Figure 3).

**Fig 3.**
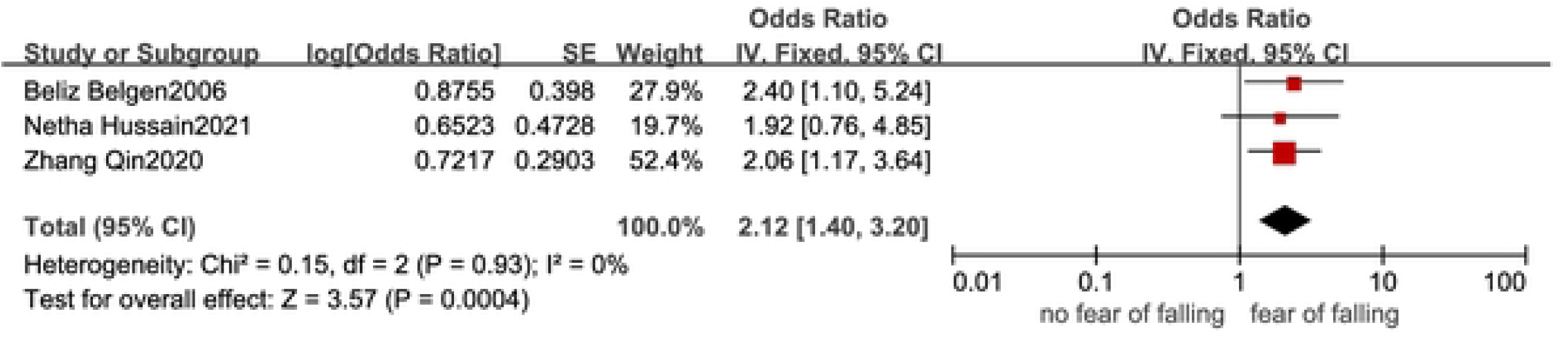
Forest plot of the relationship between history of falls and FOF.

#### 2.3.3 Balance ability

Two studies reported a correlation between balance ability and fear of falling, with heterogeneity tests (I^2^=0%, p=0.35), and there was no heterogeneity among the studies, so a fixed-effects model was used for the analysis. The results showed that balance ability(OR=3.05; 95% CI:1.60-5.80) was a risk factor for fear of falls in chronic stroke(Figure 4).

**Fig 4.**
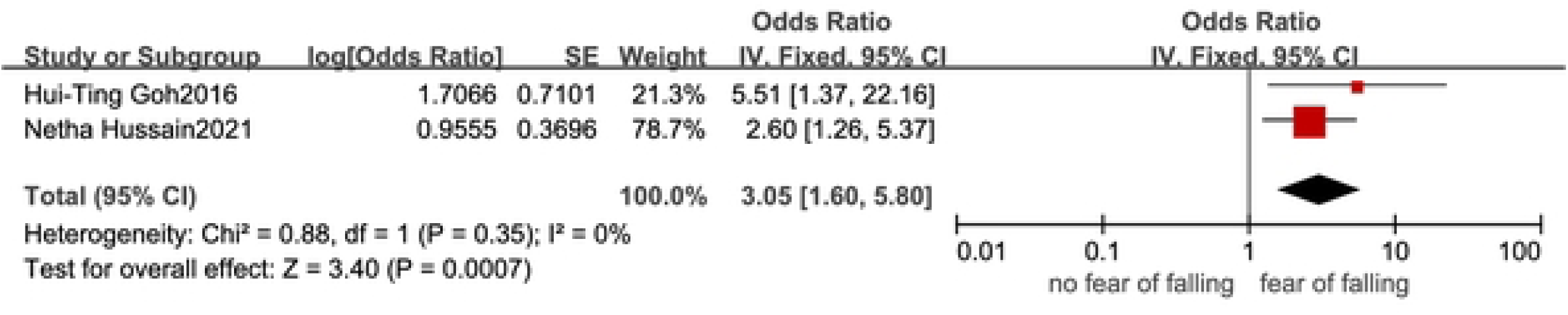
Forest plot of the relationship between balance ability and FOF.

#### 2.3.4 Mobility

Two studies reported a correlation between mobility and fear of falling, with heterogeneity testing (I^2^=87%, p=0.0005), the high heterogeneity between studies may be related to the different assessment tools used by the two studies in assessing Mobility, one study using The Functional Ambulation Category and the other using the “Stand up-GO” timing test.The results showed no statistical significance(OR=2.75;95%CI:0.29-25.63)(Figure 5).

**Fig 5.**
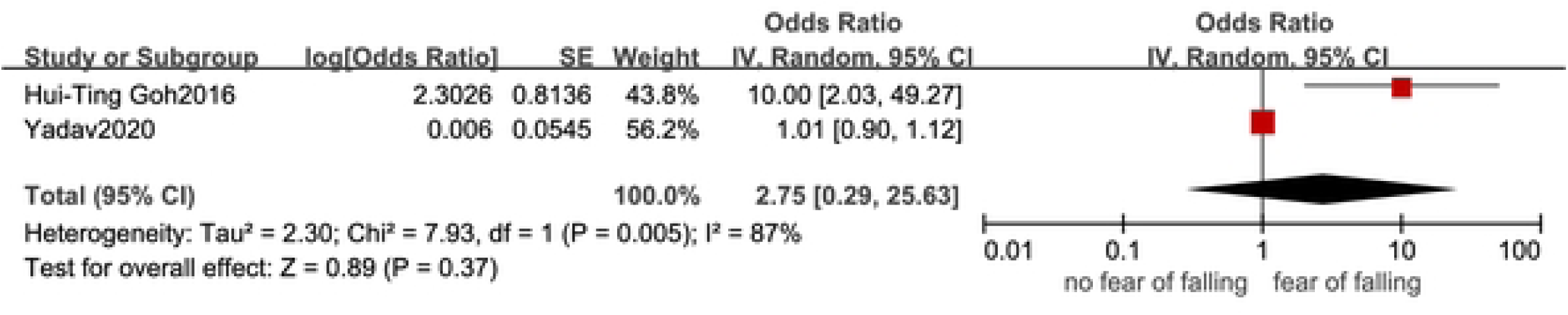
forest plot of the relationship between mobility and FOF.

#### 2.3.5 Other Factors

The following influencing factors were only mentioned in a single study, and meta-analysis could not be conducted, so descriptive analysis was used, in which anxiety (OR=2.29;95%CI: 1.43-3.67), depression (OR=1.80;95%CI:1.22-2.67), poor motor function of lower extremity (OR=1.14;95% CI:1.00-1.29), physical inactivity (OR=2.04;95%CI:1.01-4.12) was a risk factor for fall fear in patients with chronic stroke.Have a spouse (OR=0.61;95%CI:0.435-0.875) is a protective factor. Zhang Qin et al(19)have found that anxiety and depression are risk factors for fear of falling, and the marital status of a spouse is the protective factor.

### 2.4 Sensitivity analysis

These four risk factors, including age, fall history, balance ability and mobility, were not significantly different after switching models,suggesting that the results have high reliability. Results of the sensitivity analysis are shown in Table 2.

**Table 2:** Sensitivity analysis.

| Risk factor | fixed effect model |  | random effect model |  |
| --- | --- | --- | --- | --- |
|  | OR | 95%CI | OR | 95%CI |
| age | 1.00 | 0.97-1.04 | 1.13 | 0.85-1.52 |
| history of falls | 2.12 | 1.40-3.64 | 2.12 | 1.40-3.64 |
| Balance ability | 3.05 | 1.60-5.80 | 3.05 | 1.60-5.80 |
| mobility | 1.02 | 0.91-1.13 | 2.75 | 0.29-25.63 |

### 2.5 Publication bias

The number of studies included in the outcome index was less than 10, so no funnel plot was made to verify publication bias.

## 3 Discussion

### 3.1 General Factors

Two of the studies included in this paper reported the relationship between gender and fear of falling, but the results of the two studies were completely opposite, so a meta-analysis was not conducted.Both studies found no statistically significant relationship between gender and fear of falling.This is inconsistent with the findings of Suna Park et al(24), who reported that women have a higher risk of FOF when walking outdoors compared to men.Laren A et al(25) also found that women are 2.25 times more likely to experience fear of falling than men in acute stroke patients.In the elderly population, female gender has been proven to be an important influencing factor for falling fear(26–29).A study(30) found that after stroke, the level of physical activity decreases and women are more prone to FOF. There is no significant difference in the level of physical activity between men and women among stroke survivors, but the walking speed of men is correlated with the level of physical activity, and the balance ability of women is correlated with the level of physical activity.FoF is positively correlated with walking speed in stroke patients(15) and negatively correlated with balance ability, which may be the reason why women are more prone to FOF than men.However, persson et al found that men are more likely to fall than women(31), and men who are more likely to fall are less likely to have a fear of falling than women, which is a question worth considering.Further research is needed on the relationship between gender and fear of falls in chronic stroke patients, and the current number of studies is limited and the results obtained are not entirely consistent.This study found no statistical significance between age and fear of falling, but Guan et al(13) found that age may be an important factor affecting the development of fear of falling in patients with chronic stroke.Yuan Lanlan et al(32) also found that age >70 years old is a risk factor for fear of falling,First of all, the elderly will show slow movement, lower limb muscle strength, blurred vision, cognitive function decline, etc. Secondly, the ability to acquire and understand disease-related knowledge is poor, which may lead to a higher degree of fear of falling in the elderly.However, the number of studies included in this paper is limited, and the results may be unstable, so more studies are needed to verify the relationship between age and FOF in chronic stroke patients.

### 3.2 Disease-related factors

Yadav et al(23) found that lower limb motor function is an important influencing factor of fall fear,For every 1-unit increase in the lower limb Fugl-Meyer score, the risk of FOF increased by 1.36 times.This is consistent with the findings of Suna Park et al(24), who reported that lower limb strength is the most important influencing factor for FOF. Therefore, improving lower limb motor function may reduce the risk of fear of falling.Furkan et al also found that there was a significant negative correlation between FOF and upper limb Fugl-Meyer score and upper limb functional arm motor ability score (r=-0.888,r=-0.926)(33).It can be found that stroke people with poor motor function are more likely to face the fear of falling, and it is hoped that more studies will be conducted to explore whether motor function is a risk factor for the fear of falling in chronic stroke.This study found that balance ability is one of the risk factors for fear of falling in chronic stroke. This is consistent with the study results of Semra Oguz et al(34), which showed that patients with poor balance have higher fear of falling.Many studies have confirmed the negative correlation between fear of falling and balance ability(13, 27, 35, 36).This study found that the history of falls is an important risk factor for the occurrence of fear of falling in chronic stroke, and the risk of falling fear in patients with a history of falling is 2.4 times that of patients without a history of falls [20].It has been reported that fear of falling is also a risk factor for falling(37, 38). What can be known is that fear of falling and falling affect each other, forming a vicious circle. Identifying patients with high fear of falling and developing intervention measures can also reduce the occurrence of falls.A Thai study found that the prevalence of FoF in patients with a history of falling was 8.21%, and that in subjects without a history of falling was 7.80%(27). It can be seen that a history of falling is not the only predictor of fear of falling, and a high proportion of FOF will also occur in people without a history of falling(39).In addition, Netha Hussain et al(22) proposed that the risk of falling fear due to physical inactivity prior to stroke was 2.6 times that of active patients. This is consistent with the research report of Hanna et al(40), which reported that the longer the sitting time of stroke patients per day, the more likely the fall fear will occur.

### 3.3 Psychological factors

This study found that anxiety and depression are risk factors for fear of falling. Guan Q et al(13) found that depression is a risk factor related to fear of falling after chronic stroke. Schmid et al(41) also found that the fear of falling after stroke was negatively correlated with anxiety and depression scores.In the elderly population, anxiety and depression have been confirmed by a number of studies to be related to the fear of falling(42–45), while in the group of chronic stroke, firstly, because of the limited number of studies on the relationship between the two. Secondly, the presentation of results between studies is not the same, resulting in some data cannot be extracted and analyzed. All these make it impossible to combine the effect size of these two risk factors in this study, so the results of this study need to be further verified by more studies.

### 3.4 Fear of fall assessment tool

Through reading the literature, it is found that The self-made questionnaire is most commonly used to screen the prevalence of fall fear, and the Fall Efficacy Scale International and the Short Falls Efficacy Scale International are used to evaluate the severity of fall fear. Because there are many versions of the current definition of the fear of falling, there are many different scales to measure the fear of falling.The use of different assessment scales may vary the results of the studies and may increase heterogeneity across studies.More and more clinical workers gradually began to pay attention to the psychological problems of patients, and the psychological problem of falling fear will get more attention. Therefore, it is hoped that more scholars will develop a standard scale specifically for evaluating stroke fear of falling in the future.

### 3.5 Limitations and recommendations

Recognizing the dangers of fear of falling and early identification of high-risk patients with fear of falling through identified risk factors can help clinical staff develop targeted interventions to reduce the occurrence of adverse outcomes during hospitalization, reduce the length and cost of patient stay, and improve the quality of care.

Limitations of this study: (1) The number of studies included was limited, and only Chinese and English literature were included, so there was a certain selection bias; (2) Due to the small number of literatures included, some influencing factors still need further verification; (3) Most of the included studies are case-control studies and cross-sectional studies, and the causal relationship is not strong enough, so more cohort studies need to be included in the future to verify the conclusions. To sum up, this study is limited by the number and quality of included studies, and the conclusions of this study need to be verified by more large sample size, multi-center, high-quality and prospective studies.

## 4 Summary

Fear of falling can have many adverse effects on patients, such as limited activities, reduced participation in activities, increased anxiety, depression and even reduced quality of life, and increased the risk of disability and relapse. A prospective study found that a higher fear of falling may be associated with an increased rate of disability(46), so it is necessary to identify the risk factors for fall fear in patients with chronic stroke early to improve the prognosis of patients. By combining the effect size of published literature, this study found that impaired balance ability and a history of falling are risk factors for fear of falling in chronic stroke, which can provide reference for clinical staff to formulate corresponding intervention measures to reduce the occurrence of fall fear.

## Data Availability

All relevant data are within the manuscript and its Supporting Information files

## Reference

1. Katan M, Luft A. Global Burden of Stroke. Semin Neurol. 2018;38(2):208–11.

2. Zhou M, Wang H, Zeng X, Yin P, Zhu J, Chen W, et al. Mortality, morbidity, and risk factors in China and its provinces, 1990-2017: a systematic analysis for the Global Burden of Disease Study 2017. Lancet. 2019;394(10204):1145–58.

3. Walsh M, Galvin R, Horgan NF. Fall-related experiences of stroke survivors: a meta-ethnography. Disabil Rehabil. 2017;39(7):631–40.

4. Ignacio KHD, Diestro JDB, Medrano JMM, Salabi SKU, Logronio AJ, Factor SJV, et al. Depression and Anxiety after Stroke in Young Adult Filipinos. J Stroke Cerebrovasc Dis. 2022;31(2):106232.

5. Rost NS, Brodtmann A, Pase MP, van Veluw SJ, Biffi A, Duering M, et al. Post-Stroke Cognitive Impairment and Dementia. Circ Res. 2022;130(8):1252–71.

6. Whitney DG, Dutt-Mazumder A, Peterson MD, Krishnan C. Fall risk in stroke survivors: Effects of stroke plus dementia and reduced motor functional capacity. J Neurol Sci. 2019;401:95–100.

7. Xu T, Clemson L, O’Loughlin K, Lannin NA, Dean C, Koh G. Risk Factors for Falls in Community Stroke Survivors: A Systematic Review and Meta-Analysis. Arch Phys Med Rehabil. 2018;99(3):563–73.e5.

8. Mansfield A, Inness EL, McIlroy WE. Stroke. Handb Clin Neurol. 2018;159:205–28.

9. Peng M, Wu B, Wang X, Ding Y, Li Y, Cheng X. Clinical Factors Affecting the Recovery of Sensory Impairment After Cerebral Infarction: A Retrospective Study. Neurologist. 2023;28(2):117–22.

10. Zhang S, Cheng S, Zhang Z, Wang C, Wang A, Zhu W. Related risk factors associated with post-stroke fatigue: a systematic review and meta-analysis. Neurol Sci. 2021;42(4):1463–71.

11. Tinetti ME, Powell L. Fear of falling and low self-efficacy: a case of dependence in elderly persons. J Gerontol. 1993;48 Spec No:35–8.

12. Goh H-T, Nadarajah M, Hamzah NB, Varadan P, Tan MP. Falls and Fear of Falling After Stroke: A Case-Control Study. Pm&R. 2016;8(12):1173–80.

13. Guan Q, Jin L, Li Y, Han H, Zheng Y, Nie Z. Multifactor analysis for risk factors involved in the fear of falling in patients with chronic stroke from mainland China. Topics in Stroke Rehabilitation. 2015;22(5):368–73.

14. Schmid AA, Rittman M. Fear of falling: an emerging issue after stroke. Top Stroke Rehabil. 2007;14(5):46–55.

15. Schinkel-Ivy A, Inness EL, Mansfield A. Relationships between fear of falling, balance confidence, and control of balance, gait, and reactive stepping in individuals with sub-acute stroke. Gait & Posture. 2016;43:154–9.

16. Schmid AA, Arnold SE, Jones VA, Ritter MJ, Sapp SA, Van Puymbroeck M. Fear of falling in people with chronic stroke. Am J Occup Ther. 2015;69(3):6903350020.

17. Schoene D, Heller C, Aung YN, Sieber CC, Kemmler W, Freiberger E. A systematic review on the influence of fear of falling on quality of life in older people: is there a role for falls? Clin Interv Aging. 2019;14:701–19.

18. Yardley L, Smith H. A prospective study of the relationship between feared consequences of falling and avoidance of activity in community-living older people. Gerontologist. 2002;42(1):17–23.

19. Zhang Qin LY, Han Xiaojing, Zhuang Xujuan, Yang Yanfang, Wang Xia. Influencing factors of fear of falling in patients with first cerebral infarction in recovery period. Chinese Journal of Modern Nursing. 2020;26(28):3929–33.

20. Belgen B, Beninato M, Sullivan PE, Narielwalla K. The association of balance capacity and falls self-efficacy with history of falling in community-dwelling people with chronic stroke. Arch Phys Med Rehabil. 2006;87(4):554–61.

21. Goh HT, Nadarajah M, Hamzah NB, Varadan P, Tan MP. Falls and Fear of Falling After Stroke: A Case-Control Study. Pm r. 2016;8(12):1173–80.

22. Hussain N, Hansson PO, Persson CU. Prediction of fear of falling at 6 months after stroke based on 279 individuals from the Fall Study of Gothenburg. Sci Rep. 2021;11(1):13503.

23. Yadav T, Bhalerao G, Shyam AK. Factors affecting fear of falls in patients with chronic stroke. Top Stroke Rehabil. 2020;27(1):33–7.

24. Park S, Cho O-H. Fear of falling and related factors during everyday activities in patients with chronic stroke. Applied Nursing Research. 2021;62.

25. Laren A, Odqvist A, Hansson P-O, Persson CU. Fear of falling in acute stroke: The Fall Study of Gothenburg (FallsGOT). Topics in Stroke Rehabilitation. 2018;25(4):256–60.

26. Arik F, Soysal P, Capar E, Kalan U, Smith L, Trott M, et al. The association between fear of falling and orthostatic hypotension in older adults. Aging Clin Exp Res. 2021;33(12):3199–204.

27. Sitdhiraksa N, Piyamongkol P, Chaiyawat P, Chantanachai T, Ratta-Apha W, Sirikunchoat J, et al. Prevalence and Factors Associated with Fear of Falling in Community-Dwelling Thai Elderly. Gerontology. 2021;67(3):276–80.

28. Frankenthal D, Saban M, Karolinsky D, Lutski M, Sternberg S, Rasooly I, et al. Falls and fear of falling among Israeli community-dwelling older people: a cross-sectional national survey. Isr J Health Policy Res. 2021;10(1):29.

29. Simsek H, Erkoyun E, Akoz A, Ergor A, Ucku R. Falls, fear of falling and related factors in community-dwelling individuals aged 80 and over in Turkey. Australas J Ageing. 2020;39(1):e16–e23.

30. Vahlberg B, Bring A, Hellström K, Zetterberg L. Level of physical activity in men and women with chronic stroke. Physiotherapy Theory and Practice. 2019;35(10):947–55.

31. Persson CU, Kjellberg S, Lernfelt B, Westerlind E, Cruce M, Hansson PO. Risk of falling in a stroke unit after acute stroke: The Fall Study of Gothenburg (FallsGOT). Clin Rehabil. 2018;32(3):398–409.

32. Yuan Lanlan. Survey analysis and nursing strategies for fear of falling in stroke atients. Journal of Shanxi Health Vocational College. 2022;32(1).

33. Bilek F. Upper limb dysfunction increases fear of falling in people with stroke. Bangladesh Journal of Medical Science. 2021;20(3):539–42.

34. Oguz S, Demirbuken I, Kavlak B, Acar G, Yurdalan SU, Polat MG. The relationship between objective balance, perceived sense of balance, and fear of falling in stroke patients. Topics in Stroke Rehabilitation. 2017;24(7):527–32.

35. Azad A, Hassani Mehraban A, Mehrpour M, Mohammadi B. Clinical assessment of fear of falling after stroke: validity, reliability and responsiveness of the Persian version of the Fall Efficacy Scale-International. Med J Islam Repub Iran. 2014;28:131.

36. Vo MTH, Thonglor R, Moncatar TJR, Han TDT, Tejativaddhana P, Nakamura K. Fear of falling and associated factors among older adults in Southeast Asia: a systematic review. Public Health. 2022.

37. Li Y, Hou L, Zhao H, Xie R, Yi Y, Ding X. Risk factors for falls among community-dwelling older adults: A systematic review and meta-analysis. Front Med (Lausanne). 2022;9:1019094.

38. Lavedán A, Viladrosa M, Jürschik P, Botigué T, Nuín C, Masot O, et al. Fear of falling in community-dwelling older adults: A cause of falls, a consequence, or both? PLoS One. 2018;13(3):e0194967.

39. Hill KD. Fear of falling: a hidden burden with or without a history of falls. Evidence-based nursing. 2019;22(1):21.

40. Janssen H, Hanna E, Crowfoot G, Mason G, Vyslysel G, Sweetapple A, et al. Participation, fear of falling and upper limb impairment is associated with high sitting time in people with stroke. International Journal of Stroke. 2018;13(1):43.

41. Schmid AA, Van Puymbroeck M, Knies K, Spangler-Morris C, Watts K, Damush T, et al. Fear of falling among people who have sustained a stroke: a 6-month longitudinal pilot study. Am J Occup Ther. 2011;65(2):125–32.

42. Bahat Öztürk G, Kılıç C, Bozkurt ME, Karan MA. Prevalence and Associates of Fear of Falling among Community-Dwelling Older Adults. J Nutr Health Aging. 2021;25(4):433–9.

43. Badrasawi M, Hamdan M, Vanoh D, Zidan S, Alsaied T, Muhtaseb TB. Predictors of fear of falling among community-dwelling older adults: Cross-sectional study from Palestine. PLoS One. 2022;17(11):e0276967.

44. Luo Y, Miyawaki CE, Valimaki MA, Tang S, Sun H, Liu M. Symptoms of anxiety and depression predicting fall-related outcomes among older Americans: a longitudinal study. BMC Geriatr. 2022;22(1):749.

45. Rivasi G, Kenny RA, Ungar A, Romero-Ortuno R. Predictors of Incident Fear of Falling in Community-Dwelling Older Adults. J Am Med Dir Assoc. 2020;21(5):615–20.

46. Makino K, Makizako H, Doi T, Tsutsumimoto K, Hotta R, Nakakubo S, et al. Impact of fear of falling and fall history on disability incidence among older adults: Prospective cohort study. Int J Geriatr Psychiatry. 2018;33(4):658–62.

